# Mobile Phone Dependence Syndrome and its influencing factors among middle school students in Guangzhou, China: a cross-sectional study

**DOI:** 10.1101/2024.05.24.24307861

**Authors:** Chang Wang, Haiyuan Zhu, Rong Lin, Hui Liu, Jinrong Li, Minying Sun, Weiquan Lin, Qin Zhou, Bing Huang, Jierong Zhao, Yingyu Yang, Ying Li, Runquan Zhang, Qiqi Wu, Xiaomei Dong

## Abstract

**Background:** Mobile Phone Dependence Syndrome (MPDS), a kind of addiction caused by extra phone use, is characterized by impairment of physical, mental, and social functioning. This study aimed to assess the prevalence of MPDS, injury due to overfocusing on phone, and explore MPDS’s associated factors among middle school students in Guangzhou, China.

**Methods:** 1928 questionnaires were collected from April 2023 to May 2023 in Liwan and Nansha districts of Guangzhou through multi-stage cluster sampling. We used the Mobile Phone Dependence Scale for Middle School Students (MPDS Scale) and analyzed its reliability and validity. Frequency distribution, Chi-square test, fisher exact test and binary logistic regression were performed in data analysis.

**Results:** The reliability and validity of the MPDS Scale were good. The prevalence of MPDS was 10.0% and injury rate because of overfocusing on phone is 11.9%. Binary logistic regression demonstrated that gender, grade, personality, father’s parenting style, time of mobile phone use on rest days, and the most important motivation for using phone might influence occurrence of MPDS among middle school students (P<0.05).

**Conclusion:** Prevalence of MPDS among middle school students in Guangzhou was at a low level relatively. Students with MPDS had higher injury proportion than students with no MPDS. Female gender, grade of senior high school, introverted character, fathers with authoritarian parenting styles, spending 6 hours above on phone on rest days, and motivation of entertainment for using phone were associated with occurrence of MPDS. These findings can help develop measures to reduce occurrence of MPDS.

## Introduction

Mobile phones, in recent years, are rapidly gaining popularity due to their powerful features. While bringing convenience to our life, mobile phone brings us a series of negative health-related concerns, of which Mobile Phone Dependence Syndrome (MPDS) is the most concerning. Billieux et al. [1]defined mobile phone addiction (MPA) as “symptoms of deregulated use, negative impact on various aspects of daily life”. There is still no uniform definition for the psychological problems caused by excessive use of mobile phones in academic studies worldwide. However, it is a psychosomatic disorder that manifests itself primarily in the excessive use of mobile phones, which leads to obvious impairment of individual’s physiological, psychological and social functions, such as eyes hurting, anxiety, depression [2], and it can even lead to injury occurrence. For example, paying too much attention on phone can be a distraction that may result in injuries while walking and crossing the street [3]. It is also a behavioral addiction that mainly involves cognitive and emotional effects and is considered similar to Internet addiction [4–7]. Several studies used terms such as mobile phone abuse (MPA), problematic smartphone use (PSU), and mobile phone dependence syndrome (MPDS) to describe the mental health problems arising from excessive mobile phone use [2, 8]. In this study, we adopted the definition—Mobile Phone Dependence Syndrome (MPDS), which is given by Dr. Shi of the Xi’an Mental Health Center [9]. He defined that MPDS is a kind of addiction caused by the beyond-control mobile phone use, accompanied by the significant impairment of function in physical, mental and social.

Lots of studies [10–13] showed that among the mobile phone users worldwide, middle school students are undoubtedly the new mainstream group. Since middle school students are in a period of rapid psychological and physiological development, they are more vulnerable to excessive use of mobile phones than other populations and thus develop MPDS. And about measurement tool of mobile phone dependence problem, the scales that currently exist are MPPUS, MPAI, PMPUQ [14–16], etc. Considering the specific definition of mobile phone dependence and based on the foreign and domestic scales about phone dependence, Chinese scholar Xiaohui Wang [17] developed Mobile Phone Dependence Scale for Middle School Students (MPDS Scale), which filled the blank of phone dependence scale for middle school students in China and it was applied by some scholars in study [18].

An article [19] reviewed a large body of literature, which covered 12 countries or cities from Asia, Europe and Africa, such as Japan, Korea, Italy and Tunisia, and found that the prevalence of mobile phone abuse and MPDS ranged from 0%∼38%. For example, a study of Italy including 2853 objects showed that the overall prevalence of problematic mobile phone use was 6.3% [20]. Another study [21] reported that the rate of mobile phone dependence among high school students in Madrid was 20.0% (26.1% in females, 13.0% in males). In 2004, a survey covered 123 junior high schools with 747 students in South Korea [22] showed that 89.7% of the students used mobile phones and 15.7% were addicted to mobile phones. In addition, a survey in Spain of showed that 14.8% of adolescents aged 12-18 years had problems with mobile phone use [5]. The rate of mobile phone dependence among Chinese adolescents estimated by Tao et al. was 26.2% [6]. In addition, a study conducted in 2021 among senior high students in Guangzhou reported that students with mobile phone dependence account for 13.5% [23].

In recent years, mobile phones are increasingly involved in learning, social interaction and entertainment of middle school students. Overuse of mobile phone will lead to dependence on phones, causing negative effects. MPDS in adolescents has caused widespread concern in society and it is undoubtedly a public health issue that is becoming increasingly prominent. Addictive behaviors have been determined to be influenced by social determinants and cultural factors [24]. Rare research to date has reported the prevalence of mobile phone dependence among middle school students in economically developed city of China. For discovering the focus of education and promote the physical and mental health of middle school students, this study on MPDS and exploring its influencing factors of students was carried out.

## Objects and Methods

### Objects selection and sampling method

Our target objects were middle school students in Guangzhou. Middle school in this study refers to the school in China’ mainland who provides secondary education and can be divided into junior high school and senior high school. This study conducted among junior and senior high school students. The research was conducted from April to May 2023 and the objects were selected by multi-stage cluster random sampling. Our sampling procedure is shown in the following figure(fig.1)

**fig 1.**
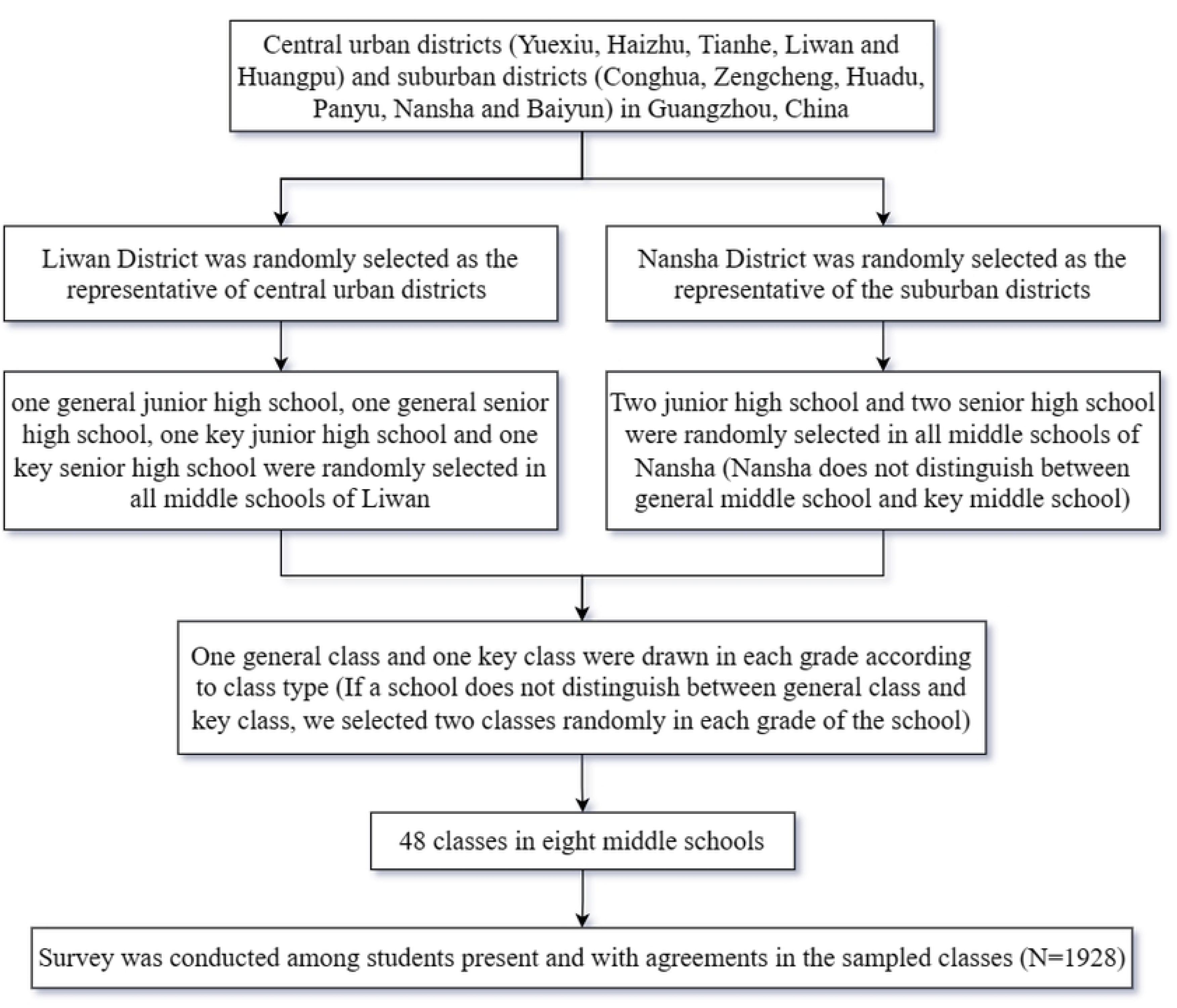
The flowchart of sampling process in this study

### Sample size estimation

In this study, based on the prevalence of middle school students’ mobile phone use dependence reported by the study of Shuman Tao [6], we set the expected prevalence of MPDS among middle school students in Guangzhou as 25.0%. Then we used the formula of simple random sampling to estimate the sample size. We set α equal to 0.05 and d equal to 0.15p.

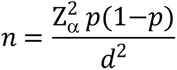

The result is that n is equal to 513. We increased the sample size by half on the basis of calculation result above, considering that our sampling method is multi-stage cluster sampling (n=770). We also took into account that the response rate is at 90% around so we figured out that the sample size of this survey should be at least 856 students.

### Research tools

#### Self-designed questionnaire

1. Personal information: including gender, age, grade, and ethnicity, etc.
2. Family information: including family residence, parental educational background, parenting styles, parental care for child, whether they are an only child.
3. Mobile phone use: including the number of mobile phones owned, time spent on mobile phone, and the amount of Internet traffic used per month, etc.
4. Consequences of mobile phone use: including aching fingers, eyes blurred or hurting, tinnitus, being injured because of overfocusing on phone (such as hitting on trees).

#### Mobile Phone Dependence Scale for Middle School Students

This scale [17] assesses students’ dependence degree on mobile phone in terms of three dimensions: abstinence, compulsivity and prominence. It consists of 16 items. Each item is rated on a five-level Likert scale ranging from “not at all consistent” to “completely consistent”. The total score for this scale is 80, and students are considered to have Mobile Phone Dependence Syndrome if their scores are over 48. The Cronbach’s coefficient of this scale in this study was 0.88.

The three dimensions of the scale are defined as follows:

1. Abstinence refers to the emotional reactions that occurs when one is suddenly forced to be unable to use his phone or is unable to use it normally, such as anxiety, irritability, etc.
2. Compulsivity refers to the fact that a person has a strong desire and urge to use the mobile phone and is difficult to extricate himself, or even has a delusion.
3. Prominence means that the behavior of individual is dominated by the use of mobile phone, which mainly manifested as phone overuse and checking phone excessively in daily life.

#### Research content

Xiaohui Wang’s Mobile Phone Dependence Scale for Middle School Students (MPDS Scale) may have different factor structures in different regions, as well as in different sample. Therefore, this study validated the reliability and validity of the scale at first.

The scores of MPDS Scale of participating students were described using means and median. The prevalence of MPDS and injury caused by overfocusing on phone was revealed through statistical description, and the influencing factors of MPDS were investigated through univariate analysis and multivariate analysis.

#### Quality control

A pre-survey of the target population was initiated before the survey began for modifying the self-designed questionnaire, and the investigators were trained in a uniform manner. The strong support of the related personnel of the school was sought to ensure the compliance of objects and the authenticity of the survey data. After finishing, investigators checked the questionnaire one by one in the classrooms. When missing items or logical errors were found, investigators would ask students to supplement or correct them immediately. During the process of data analysis, our exclusion criteria is: (1) Students who did not complete Mobile Phone Dependence Scale for Middle School Students (one item missing is allowed); (2) Students who did not answer the multiple-choice questions in a logical and serious manner.

### Statistical Methods

#### Scale reliability and validity test

The reliability coefficient test was performed using SPSS 26.0, while the sample was subjected to confirmatory factor analysis using Amos 26.0.

#### Statistical analyses

SPSS 26.0 was used for data analysis. Objects with one missing item of MPDS Scale in the validated questionnaire were interpolated using mean values. We compared the distribution of MPDS between participants with different characteristics, as well as consequences of mobile phone use in MPDS group and non-MPDS group by chi-square or fisher exact test. Binary logistic regression model was used to identify factors associated MPDS, and the independent variables included in the model were identified based on relevant studies and univariate analysis. The odds ratio (*OR*) and 95% confidence intervals (95% CI) were used to estimate the effects of the independent variables. The significant level was 0.05(two-tailed).

### Ethical considerations

Ethical approval for the current study was obtained from Ethics Committee of Guangzhou Center for Disease Control and Prevention, China (approval number: GZCDC-ECHR-2024P0095). This study has obtained verbal consent from the participants and their guardians. The researchers had informed the participants (and their legal guardian) of the research background, purpose, steps, risks and benefits of the project. They had enough time to read the informed consent and discuss with others and they had the rights to get the answers of their any questions about the study from researchers. After ensuring that the participant fully understood all aspects of the study, they verbally expressed their willingness to participate by stating,“**I consent**.”and then they had right to decide whether to fill out the questionnaire.The participants (or legal guardian) were able to quit from the study if they wanted at any time.

## Results

### Basic information of survey respondents

A total of 1928 questionnaires were collected, of which 1883 questionnaires were defined valid. The effective rate was 97.67%. The mean age of sample was 15.33 years (*SD*=1.71 years). Our study found that the prevalence of MPDS among middle school students in Guangzhou was 10.0%.

### Reliability and validity test of Mobile Phone Dependence Scale for Middle School Students (N=1883)

The Cronbach’s coefficient of the MPDS Scale of sample is 0.880 and the split-half reliability is 0.859, which indicated that the reliability of scale is good. Since the MPDS Scale is a well-established scale, a confirmatory factor analysis can be conducted for the original.

We used Amos.26 to conduct confirmatory factor analysis on the study sample. The fit indices of the MPDS Scale meet the index standard (online supplemental table 1), indicating that the scale is suitable for sample of our study.

## Scores of MPDS Scale among middle school students in Guangzhou (N=1883)

The mean score of MPDS Scale of the participants was 35.74 and the median was 35. No matter in all participants or the participants with MPDS, the mean score for each item of prominence was the highest, followed by abstinence and then compulsivity.

### The comparison of consequences of mobile phone use

The results showed that there were differences in the consequences of phone use between the two groups (table 2). The rate of being injured due to overfocusing on phone in MPDS group is 29.4%, significantly higher than another group (10%). The findings proved that excessive attention to mobile phone not only causes adverse physical effects, but also may lead to injuries, such as hitting a tree and fall.

**Table 1.** Scores of each item at different dimensions of MPDS Scale.

| Scales and Dimensions | All participants |  |  | Participants with MPDS |  |  |
| --- | --- | --- | --- | --- | --- | --- |
|  | MEAN | SD | MEDIAN | MEAN | SD | MEDIAN |
| Abstinence | 2.12 | 0.88 | 2.00 | 3.64 | 0.64 | 3.50 |
| Compulsivity | 1.71 | 0.71 | 1.60 | 2.77 | 0.80 | 2.80 |
| Prominence | 2.90 | 0.80 | 3.00 | 3.89 | 0.50 | 3.80 |
| Total Score | 35.74 | 10.61 | 35.00 | 55.06 | 5.90 | 54.00 |

**Table 2.** Comparison of consequences of mobile phone use among middle school students in Guangzhou (n/%)

| Consequences of mobile phone use | Total n *(%) | Non-MPDS group | MPDS group | $\chi^2$ | P value |
| --- | --- | --- | --- | --- | --- |
| Aching fingers |  |  |  |  |  |
| Yes | 227(12.1) | 172(10.2) | 55(29.4) | 58.323 | <0.001 |
| No | 1646(87.9) | 1514(89.8) | 132(70.6) |  |  |
| Eyes blurred or hurting |  |  |  |  |  |
| Yes | 630(33.6) | 522(30.9) | 108(57.8) | 54.318 | <0.001 |
| No | 1245(66.4) | 1166(69.1) | 79(42.2) |  |  |
| Tinnitus |  |  |  |  |  |
| Yes | 150(8.0) | 116(6.9) | 34(18.2) | 29.258 | <0.001 |
| No | 1725(92.0) | 1572(93.1) | 153(81.8) |  |  |
| Being injured because of overfocusing on phone |  |  |  |  |  |
| Yes | 223(11.9) | 168(10.0) | 55(29.4) | 60.423 | <0.001 |
| No | 1646(88.1) | 1514(90.0) | 132(70.6) |  |  |

\*There are a little data missing

### Univariate analysis of MPDS among middle school students in Guangzhou (N=1883))

We found in our study that the prevalence of MPDS is 10.0%. The results (table 3) showed that in the individual characteristics, there were differences in the distribution of MPDS among middle school students in genders, grades, personalities, evaluation of current study pressure and academic achievements (*P*<0.05).

**Table 3.**
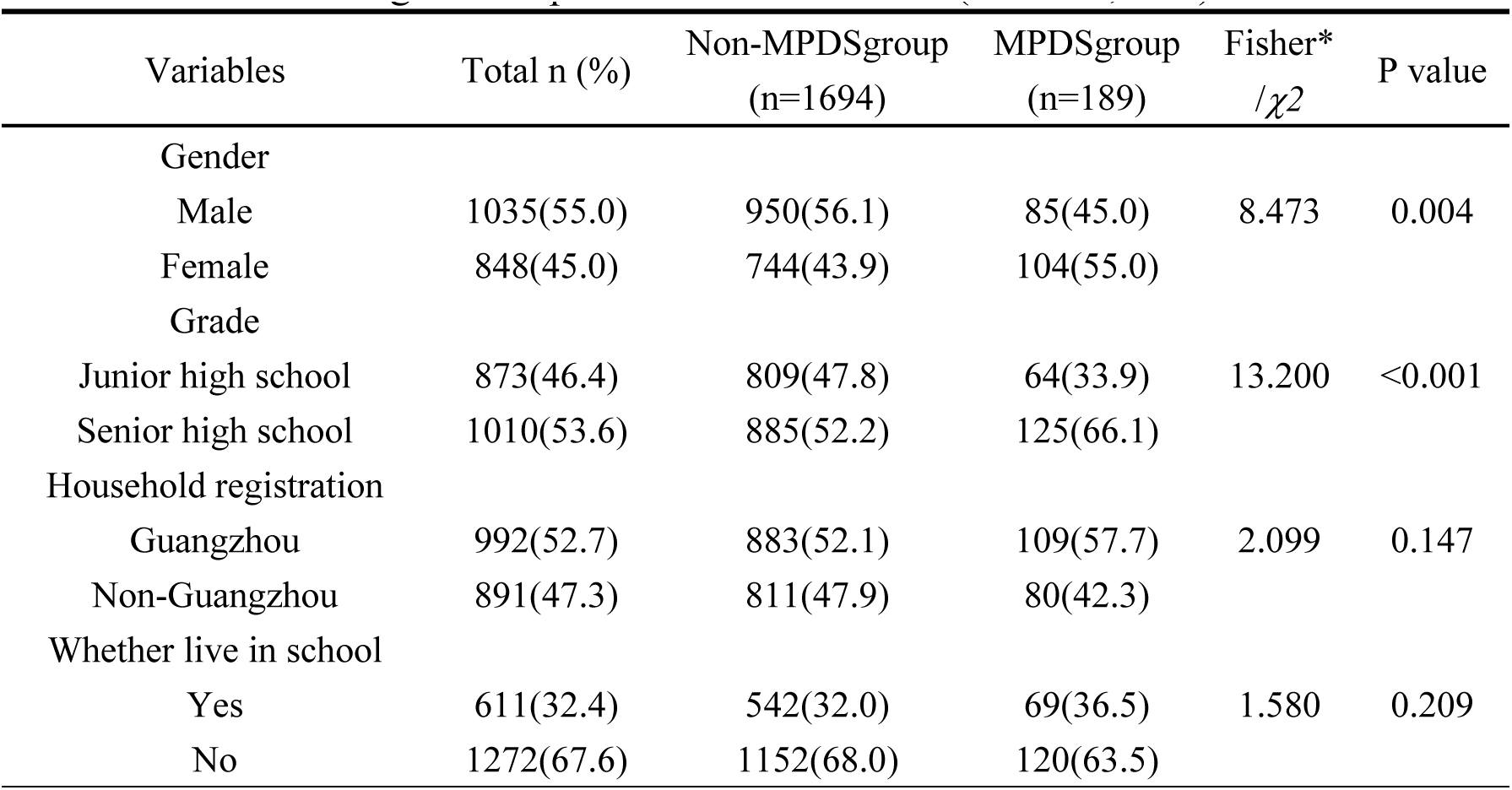
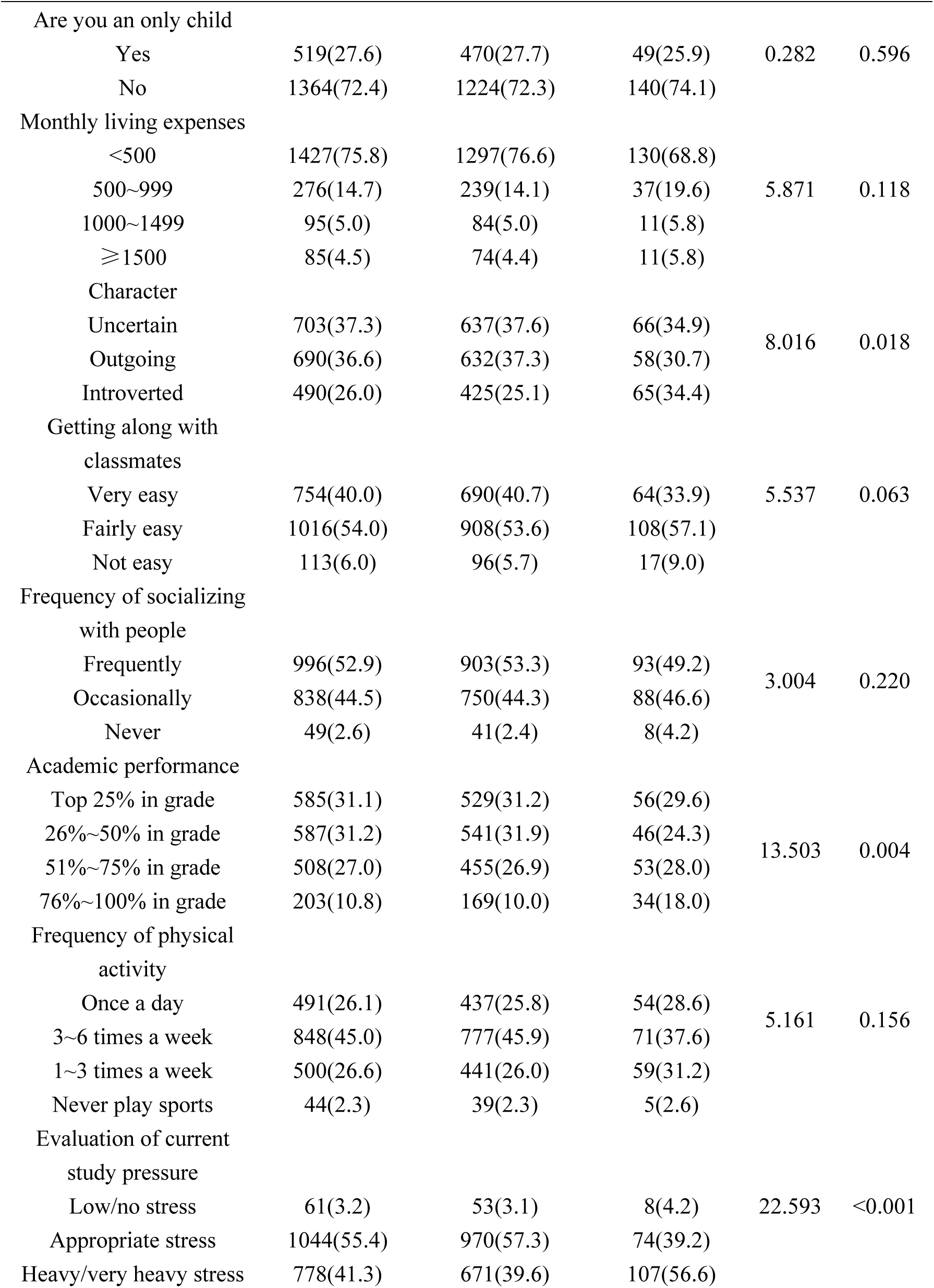
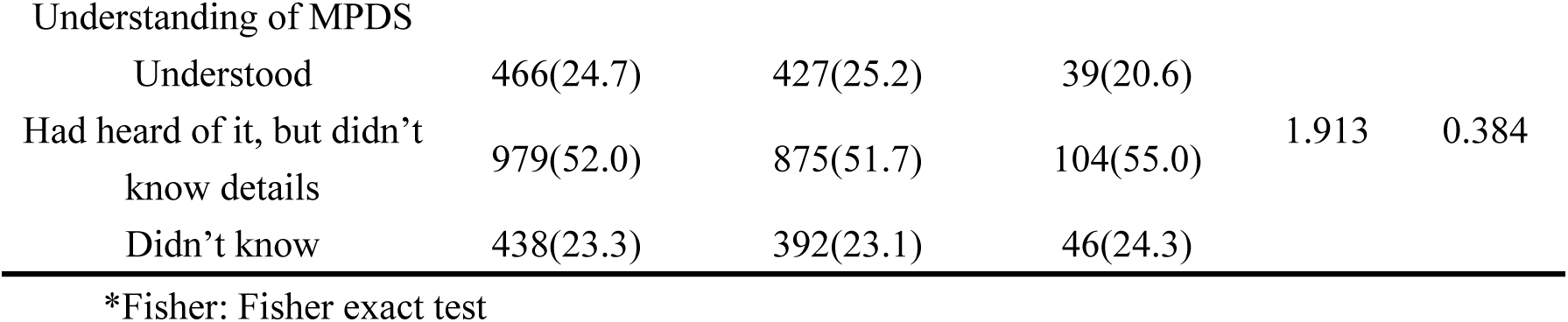
Distribution and comparison of MPDS among middle school students in Guangzhou of personal characteristics (N=1883; n/%)

| Variables | Total n (%) | Non-MPDSgroup<br>(n=1694) | MPDSgroup<br>(n=189) | Fisher*<br>/$\chi^2$ | P value |
| --- | --- | --- | --- | --- | --- |
| Gender |  |  |  |  |  |
| Male | 1035(55.0) | 950(56.1) | 85(45.0) | 8.473 | 0.004 |
| Female | 848(45.0) | 744(43.9) | 104(55.0) |  |  |
| Grade |  |  |  |  |  |
| Junior high school | 873(46.4) | 809(47.8) | 64(33.9) | 13.200 | <0.001 |
| Senior high school | 1010(53.6) | 885(52.2) | 125(66.1) |  |  |
| Household registration |  |  |  |  |  |
| Guangzhou | 992(52.7) | 883(52.1) | 109(57.7) | 2.099 | 0.147 |
| Non-Guangzhou | 891(47.3) | 811(47.9) | 80(42.3) |  |  |
| Whether live in school |  |  |  |  |  |
| Yes | 611(32.4) | 542(32.0) | 69(36.5) | 1.580 | 0.209 |
| No | 1272(67.6) | 1152(68.0) | 120(63.5) |  |  |
| Are you an only child |  |  |  |  |  |
| Yes | 519(27.6) | 470(27.7) | 49(25.9) | 0.282 | 0.596 |
| No | 1364(72.4) | 1224(72.3) | 140(74.1) |  |  |
| Monthly living expenses |  |  |  |  |  |
| <500 | 1427(75.8) | 1297(76.6) | 130(68.8) |  |  |
| 500~999 | 276(14.7) | 239(14.1) | 37(19.6) | 5.871 | 0.118 |
| 1000~1499 | 95(5.0) | 84(5.0) | 11(5.8) |  |  |
| ≥1500 | 85(4.5) | 74(4.4) | 11(5.8) |  |  |
| Character |  |  |  |  |  |
| Uncertain | 703(37.3) | 637(37.6) | 66(34.9) | 8.016 | 0.018 |
| Outgoing | 690(36.6) | 632(37.3) | 58(30.7) |  |  |
| Introverted | 490(26.0) | 425(25.1) | 65(34.4) |  |  |
| Getting along with classmates |  |  |  |  |  |
| Very easy | 754(40.0) | 690(40.7) | 64(33.9) | 5.537 | 0.063 |
| Fairly easy | 1016(54.0) | 908(53.6) | 108(57.1) |  |  |
| Not easy | 113(6.0) | 96(5.7) | 17(9.0) |  |  |
| Frequency of socializing with people |  |  |  |  |  |
| Frequently | 996(52.9) | 903(53.3) | 93(49.2) | 3.004 | 0.220 |
| Occasionally | 838(44.5) | 750(44.3) | 88(46.6) |  |  |
| Never | 49(2.6) | 41(2.4) | 8(4.2) |  |  |
| Academic performance |  |  |  |  |  |
| Top 25% in grade | 585(31.1) | 529(31.2) | 56(29.6) |  |  |
| 26%~50% in grade | 587(31.2) | 541(31.9) | 46(24.3) | 13.503 | 0.004 |
| 51%~75% in grade | 508(27.0) | 455(26.9) | 53(28.0) |  |  |
| 76%~100% in grade | 203(10.8) | 169(10.0) | 34(18.0) |  |  |
| Frequency of physical activity |  |  |  |  |  |
| Once a day | 491(26.1) | 437(25.8) | 54(28.6) | 5.161 | 0.156 |
| 3~6 times a week | 848(45.0) | 777(45.9) | 71(37.6) |  |  |
| 1~3 times a week | 500(26.6) | 441(26.0) | 59(31.2) |  |  |
| Never play sports | 44(2.3) | 39(2.3) | 5(2.6) |  |  |
| Evaluation of current study pressure |  |  |  |  |  |
| Low/no stress | 61(3.2) | 53(3.1) | 8(4.2) | 22.593 | <0.001 |
| Appropriate stress | 1044(55.4) | 970(57.3) | 74(39.2) |  |  |
| Heavy/very heavy stress | 778(41.3) | 671(39.6) | 107(56.6) |  |  |
| Understanding of MPDS |  |  |  |  |  |
| Understood | 466(24.7) | 427(25.2) | 39(20.6) | 1.913 | 0.384 |
| Had heard of it, but didn't know details | 979(52.0) | 875(51.7) | 104(55.0) |  |  |
| Didn't know | 438(23.3) | 392(23.1) | 46(24.3) |  |  |

\*Fisher: Fisher exact test

In the comparison of family conditions among MPDS group and non-MPDS group (table 4), differences were found in fathers’ levels of education, parenting styles and degrees of care for their children(*P*<0.05).

**Table 4.** Distribution and comparison of MPDS among middle school students in Guangzhou with different family conditions(N=1883; n/%)

| Variables | Total n (%) | Non-MPDS group<br>(n=1694) | MPDSgroup<br>(n=189) | Fisher<br>* /$\chi^2$ | P value |
| --- | --- | --- | --- | --- | --- |
| Family residence |  |  |  |  |  |
| Rural area/Town | 725(38.5) | 646(38.1) | 79(41.8) | 0.964 | 0.326 |
| City | 1158(61.5) | 1048(61.9) | 110(58.2) |  |  |
| Father's Education |  |  |  |  |  |
| Elementary school or below | 100(5.3) | 84(5.0) | 16(8.5) | 10.497 | 0.033 |
| Junior high school | 668(35.5) | 597(35.2) | 71(37.6) |  |  |
| Senior high/ Technical secondary school | 636(33.8) | 588(34.7) | 48(25.4) |  |  |
| Junior college/ Vocational university | 253(13.4) | 221(13.2) | 32(16.9) |  |  |
| Bachelor's degree or above | 226(12.0) | 204(12.0) | 22(11.6) |  |  |
| Mother's education |  |  |  |  |  |
| Elementary school or below | 193(10.2) | 175(10.3) | 18(9.5) | 6.784 | 0.148 |
| Junior high school | 737(39.1) | 666(39.3) | 71(37.6) |  |  |
| Senior high / Technical secondary school | 519(27.6) | 464(27.4) | 55(29.1) |  |  |
| Junior college/Vocational university | 233(12.4) | 201(11.9) | 32(16.9) |  |  |
| Bachelor's degree or above | 201(10.7) | 188(11.1) | 13(6.9) |  |  |
| Father's parenting style |  |  |  |  |  |
| Democratic | 1023(54.3) | 947(55.9) | 76(40.2) |  |  |
| Authoritarian | 401(21.3) | 343(20.2) | 58(30.7) | 18.413 | <0.001 |
| Indulgent | 442(23.5) | 389(23.0) | 53(28.0) |  |  |
| Spoiling | 17(0.9) | 15(0.9) | 2(1.1) |  |  |
| Mother's parenting style |  |  |  |  |  |
| Democratic | 1083(57.5) | 989(58.4) | 94(49.7) |  |  |
| Authoritarian | 372(19.8) | 329(19.4) | 43(22.8) | 5.684 | 0.118 |
| Indulgent | 405(21.5) | 356(21.0) | 49(25.9) |  |  |
| Spoiling | 23(1.2) | 20(1.2) | 3(1.6) |  |  |
| Father's caring level |  |  |  |  |  |
| Very caring | 935(49.7) | 864(51.0) | 71(37.6) |  |  |
| Average | 807(42.9) | 708(41.8) | 99(52.4) | 12.475 | 0.002 |
| Doesn't care | 141(7.5) | 122(7.2) | 19(10.1) |  |  |
| Mother's caring level |  |  |  |  |  |
| Very caring | 1257(66.8) | 1138(67.2) | 119(63.0) |  |  |
| Average | 541(28.7) | 478(28.2) | 63(33.3) | 2.307 | 0.315 |
| Not caring | 85(4.5) | 78(4.6) | 7(3.7) |  |  |

\*Fisher: Fisher exact test

In terms of mobile phone use (table 5), numbers of mobile phones owned, years of mobile phone ownership, daily mobile phone use time, the motivation and application of using mobile phone, and monthly Internet traffic were found to be different between MPDS group and non-MPDS group(*P*<0.05).

**Table 5.**
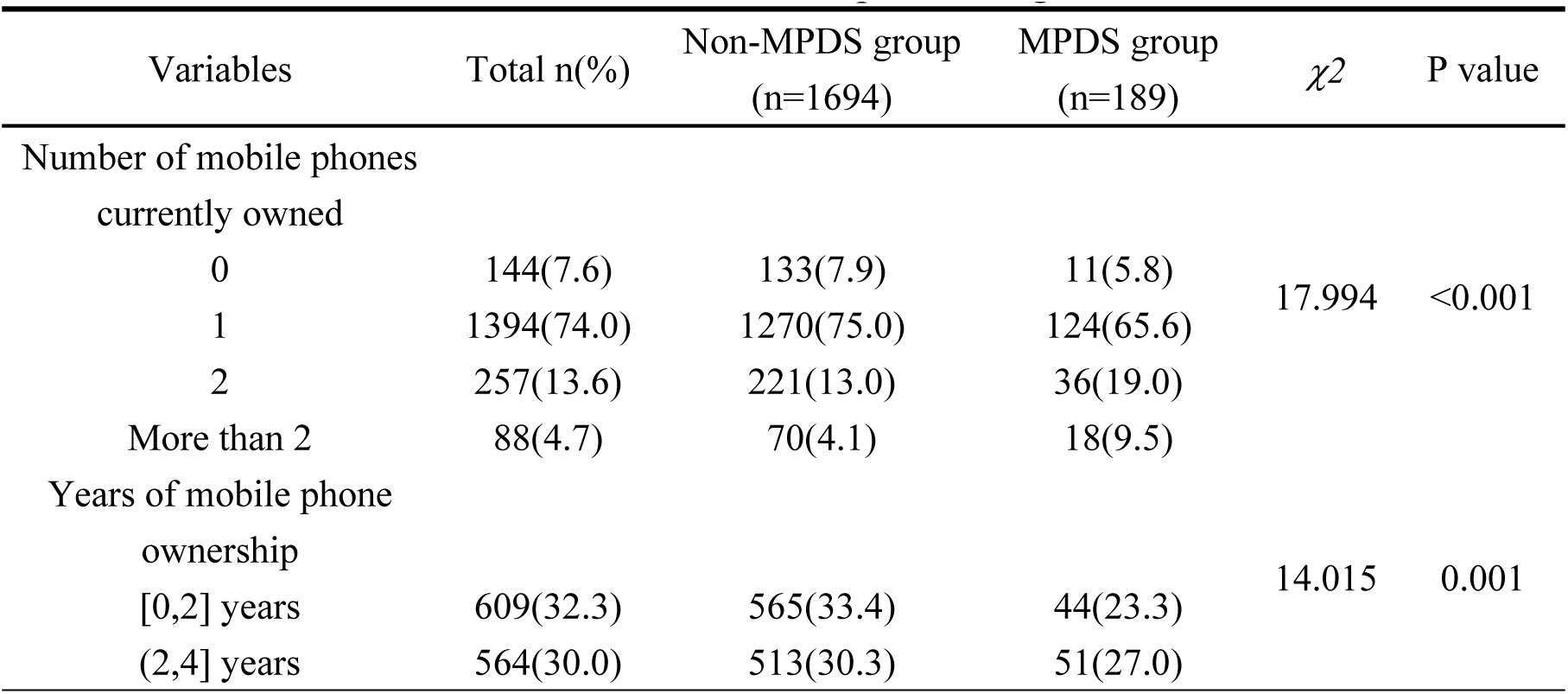
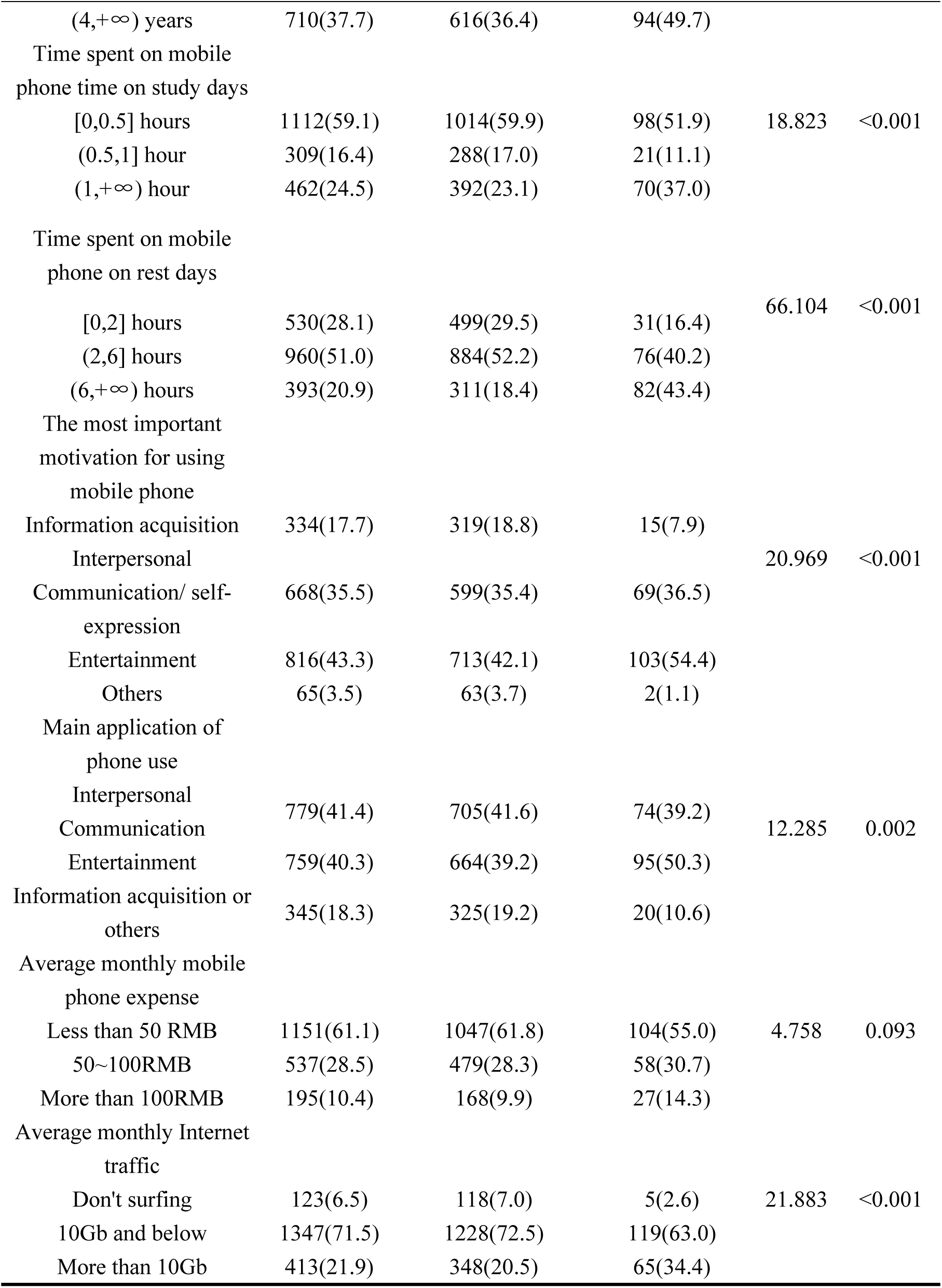
Distribution and comparison of MPDS among middle school students with different mobile phone usages.

### 3.6 Multivariate binary logistic analysis of MPDS among middle school students in Guangzhou (N=1883)

We set the “Non-MPDS” group as the reference group and variables whose P-value less than 0.05 in the univariate analysis were included to establish a binary logistic regression model. Omnibus test of the model coefficients was significant (*P*<0.001). The result of the Hosmer Lemercio test showed that the binary logistic model is well fitted to observations (*P*>0.05).

Binary logistic regression (table 6) showed that the associated factors for developing MPDS were girl (adjusted *OR*=1.772,95%*CI*:1.239-2.532), grade of high school (*aOR*=1.479, 95%*CI*:1.014-2.155), introverted personality (*aOR*=1.686, 95%*CI*:1.108-2.566), father’s authoritarian parenting styles (*aOR*=2.023, 95%*CI*:1.315-3.111), spending 6 hours above on phone on rest days (*aOR*=3.115, 95%*CI*:1.829-5.307), motivation of interpersonal communication or self-expression (*aOR*=2.197, 95%*CI*:1.183-4.079), and motivation of entertainment (*aOR*=2.527, 95%*CI*:1.381-4.624).

**Table 6.**
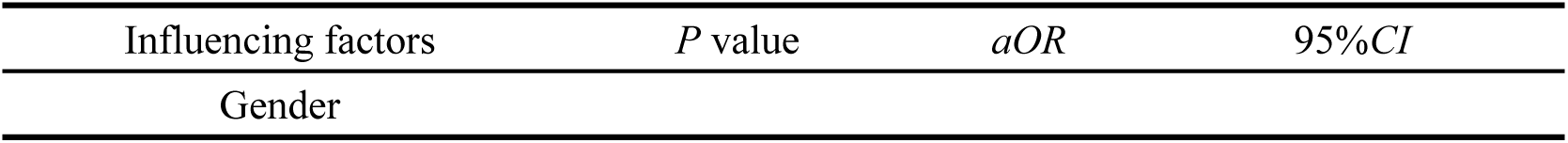
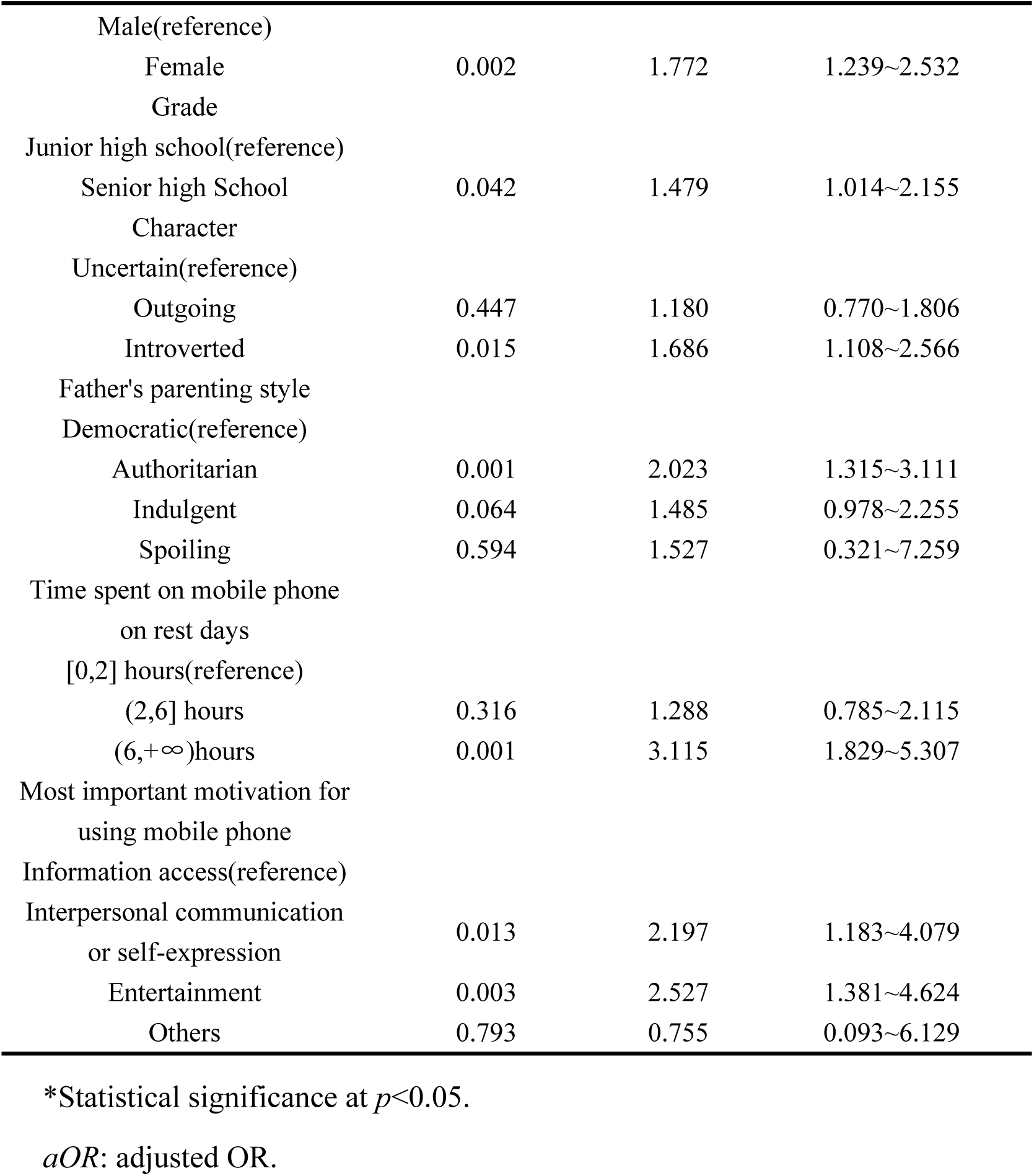
Multivariate binary logistic regression analysis of MPDS among middle school students in Guangzhou, China.

## Discussion

In MPDS Scale, we found that the mean score for each item of prominence was highest (2.90) while compulsivity was the lowest (1.71). Among the 189 students with MPDS, the highest mean score for each item was still prominence (3.89) and compulsivity stayed lowest (2.77), which indicated that principal manifestations of the syndrome of students with MPDS were negative emotional reactions when they are unable to use phone and daily behavior dominated by phone use, such as phone overuse and excessive checking of phone.

We calculated that the prevalence of MPDS in our study was 10.0%, which was similar to the results of studies in Korea, Spain and Belgium[25–27]. Our results showed that prevalence of being injured caused by overfocusing on mobile phone was 11.9% and it was 29.4% in students with MPDS. Some surveys showed that 10.7% of teenagers reported being struck or nearly struck by a vehicle because of talking on the phone and 7.7% reported texting [28]. As the number of mobile phones in use increases and the time spent on them increases, the numbers of injuries may also increase [29]. These findings suggest that the problem of mobile phone overuse among adolescents need urgent and appropriate intervention.

Similar to the results of us, other studies had reported that females are more likely to develop MPDS than males (*aOR* = 1.772)[30–32], which may be related to the fact that adolescent girls need more emotional satisfaction in social relationships than males and are more inclined to express their feelings through text messaging via mobile phone chatting applications. This suggests that school and family should strengthen emotional support and care for female middle school students

Compared to junior high school students, senior high school students were more likely to develop MPDS (*aOR* = 1.479). A survey on mobile phone dependence for middle school students in Xiamen, China, showed that high school students had higher dependence on mobile phones[33]. A possible explanation for this could be that Chinese senior high school students get heavy study burden and face a lot of pressure from entrance exam for university. Senior high school students are more likely to have difficulty in controlling their mobile phone use[33], and may use their mobile phones to relieve burnout. Therefore, it is advisable for senior high schools to provide psychological counseling regularly and alleviate academic burden for students, empowering them to deal with study pressure and problems of mobile phone dependence.

The results showed that introverted students had a higher risk of developing MPDS than students with uncertain personalities (*aOR* = 1.686). Students with ‘uncertain personality’ might be in the middle status between introversion and extroversion so that they were not sure whether they are introverted or extroverted. Middle school students with introverted personality may have been out of real-life interactions for a long time and lack the opportunity to interact with people face to face[34]. Some studies have indicated that both introverts or extroverts might be addicted to mobile phone, but introverts are more likely to use mobile phones as a programming tool and instrumental means, while extroverts use mobile phones more for socialization[35].

Our study found that students whose father with authoritarian parenting styles had a higher risk of developing MPDS (*aOR* = 2.023). A study in Indonesia found that authoritative parenting style is positively associated with mobile phone dependence: the more authoritarian parenting style parents are, the more likely mobile phone dependence children develop [36]. Interaction of parent-child cohesion and parent-child relationship quality is negatively related to mobile phone dependence [37]. In addition, a study on the correlation between family cohesion and mobile phone addiction showed that family cohesion was negatively correlated with mobile phone dependence [38] and individuals who grow up in a harmonious family and feel more emotional warmth will develop secure attachment for family, which reduces the likelihood of developing mobile phone dependence [39]. Therefore, the control of MPDS can start from family environment, such as, specifically, creating a democratic family atmosphere and communicating more with children.

In our study, spending 6 hours above on phone on rest days (*aOR* = 3.115) and mobile phone use motivated by interpersonal communication or self-expression (*aOR* = 2.197) and entertainment (*aOR* = 2.527) may be risk factors for MPDS. In rest days ( weekends and holidays), students have more disposable time, may increase the risk of excessive mobile phone use. Related studies show that there is a significant positive correlation between mobile phone use motivation and mobile phone dependence [40]. Smartphones meet people’s need for entertainment and socialization, but they may have the risk of being overused [41, 42], which may also increase the duration of mobile phone use. Middle school students should establish a correct view of mobile phone and realize that phone is also an effective learning tool.

This study has the following limitations. Firstly, sample of this study was from eight middle schools in Liwan and Nansha districts of Guangzhou. Although it was a multicenter study, the sample size was not large, for which reason that potential influencing factors may not be identified. Secondly, this study was a cross-sectional study and it was unable to confirm causal relationship between different factors and MPDS. More longitudinal studies are needed in future.

In any case, this study revealed recent prevalence of MPDS and being injured caused by overfocusing on phone of middle school students in Guangzhou, China. And we found some associated factors contributing to developing MPDS among them.

## Data Availability

The data are held or will be held in a public repository

## Acknowledgments

The authors thank all the participating students and their cooperation. Sincerely thank the officials of middle schools we surveyed for their great support. The authors also thank experts who gave advice on the revision of the questionnaire and all the investigators who collected the original data in eight middle schools for their hard work.

## Contributors

CW and HZ were responsible for the data analysis and were the major contributor for writing the manuscript. RL, HL, JL, MS, WL and QZ were responsible for research design, questionnaire design and quality control. BH, JZ, YY and YL participated in the field investigation, data collation and enter. RZ and QW participated in partial writing of the manuscript. XD gave valuable advice for the revision of the whole article.

## Ethics approval

This study was reviewed and approved by the Ethics Committee of Guangzhou Center for Disease Control and Prevention, Guangzhou, China (approval number GZCDC-ECHR-2024P0095). Informed consent was obtained from all the student participants.

## Data availability

All relevant data are within the paper and Supporting Information files.

